# Global lessons and Potential strategies in combating COVID-19 pandemic in Ethiopia: Systematic Review

**DOI:** 10.1101/2020.05.23.20111062

**Authors:** Yimam Getaneh, Ajanaw Yizengaw, Sisasy Adane, Kidist Zealiyas, Zelalem Abate, Sileshi Leulseged, Hailemichael Desalegn, Getnet Yimer, Ebba Abate

## Abstract

**Background:** Coronavirus disease 2019 (COVID-19) is a rapidly emerging disease that has been classified a pandemic by the World Health Organization (WHO). In the absence of treatment for this virus, there is an urgent need to find alternative public health strategies to control the spread. Here, we have conducted an online search for all relevant public health interventions for COVID-19. We then characterize and summarize the global COVID-19 pandemic situation and recommend potential mitigation strategies in the context of Ethiopia.

**Methods:** Initial search of Pub Med central and Google scholar was undertaken followed by analysis of the text words; COVID-19,SARS-CoV-2, Global lessons and Pandemic; A second search using all identified keywords including COVID-19, Epidemiology, Sociocultural, Ethiopia; thirdly, the reference list of all identified reports and articles were searched. Accordingly, of the 1,402 articles, 39 were included in the analysis for this review.

**Result:** Countries COVID-19 mitigation strategies widely varied. The most common global COVID-19 mitigation strategies include; whole of government approach including individual, community and environmental measures, detecting and isolating cases, contact tracing and quarantine, social and physical distancing measures including for mass gatherings and international travel measures. Models revealed that, social and physical distancing alone could prevent the pandemic from 60-95%, if timely and effectively implemented. Moreover, detecting and isolation of cases were found to be crucial while access to testing was found to the global challenge. Individual measures including proper hand washing were also reported to be effective measures in preventing the pandemic. Asymptomatic cases of COVID-19 ranged from 25% to 80% and as a result, countries are revising the case definition for early detection of mild symptomatic cases of COVID-19 with inclusion of Chills, Muscle pain and new loss of taste or smell in addition to Cough, Shortness of breath, Fever and Sore throat. Global reports also revealed that the incubation period of COVID-19 could go to 24 days. Ethiopia is also unique in the aspects of sociocultural prospects while more than 99.3% of the population has a religion. Moreover, 69% of the population is under the age of 29 years old and the health policy in the country focused on prevention and primary health care. All these could be potential entries and opportunities to combat COVID-19 pandemic in the context of Ethiopia.

**Conclusion:** While recommendations may change depending on the level of outbreak, we conclude that in Most countries have benefited from early interventions and in setups like Africa including Ethiopia where health system capability is limited, community engagement supported by local evidence with strict implementation of social and physical distancing measures is mandatory. Active involvement of religious Institutions and mobilizing youth could be entry to increase public awareness in mitigating COVID-19. Community level case detection could enhance early identification of cases which could be implemented through the health extension program. Isolation and quarantine beyond 14 days could help identify long term carriers of COVID-19. Validation and use of rapid test kits could be vital to increase access for testing. Revision of case definitions for COVID-19 could be important for early detection and identification of mild symptomatic cases.

## 1. Introduction

### 1.1. Background

On 30^th^ January 2020, the WHO declared the Chinese outbreak of COVID-19 to be a Public Health Emergency of International Concern posing a high risk to countries with vulnerable health systems^5^. The emergency committees have stated that the spread of COVID-19 may be interrupted by early detection, isolation, prompt treatment, and the implementation of a robust system to trace contacts^6^. Other strategic objectives include a means of ascertaining clinical severity, the extent of transmission, and optimizing treatment options. A key goal is to minimize the economic impact of the virus and to counter misinformation on a global scale. In light of this, various bodies have committed to making articles pertaining to COVID-19 immediately available via open access to support a unified global response^7^.

### 1.2. Global Epidemiology of COVI-19

According to the Health Commission of Hubei province, China, on December 31-2019, although 27 patients were initially announced to be afflicted with this mysterious disease, the number rose to 41 with seven critically ill patients; one death was noted in the subsequent report on January 11, 2020. The Chinese authorities reporting to WHO stated that some of the patients were operating dealers or vendors in the Huanan seafood market, which was subsequently reported to be selling live and freshly slaughtered hunted animals^8^.

Several reports of clusters of cases among families and infection of 16 health care workers pointed to human-to-human transmission of the virus. Despite recognition of the outbreak within a few weeks by the Chinese using their efficient surveillance network and laboratory infrastructure, efforts to prevent the spread of this virus were not sufficient; as of February 3, 2020, at least 17, 496 cases with a death toll of 362 in more than 25 countries have been reported^9^. In a study of 425 cases infected up to January 4, the basic reproductive number, or R0, of the virus was estimated to be approximately 2,226. This means that each patient can, on average, spread the infection to more than two healthy persons^12^.

Presence of cases with mild clinical presentation and lack of infrastructure to provide isolation for infected individuals and their close contacts, particularly in resource-limited countries, are nonetheless all hurdles to control this infection. Furthermore, based on experience, proper precautionary measures to prevent nosocomial transmission of the virus is mandatory; the majority of patients with SARS-CoV and MERS-CoV-2 had become infected in health care settings. Considering the plethora of comorbid conditions present in hospital populations, dire complications could arise in the setting of an outbreak^10^.

As of 14, April-2020 the total number of death was reported to be 6,839 while people in the above 75 accounted the share of death 47.7% (Table-1). Men are highly affected than female which accounted 61.8% of the total death (Table-2).

**Table-1:**
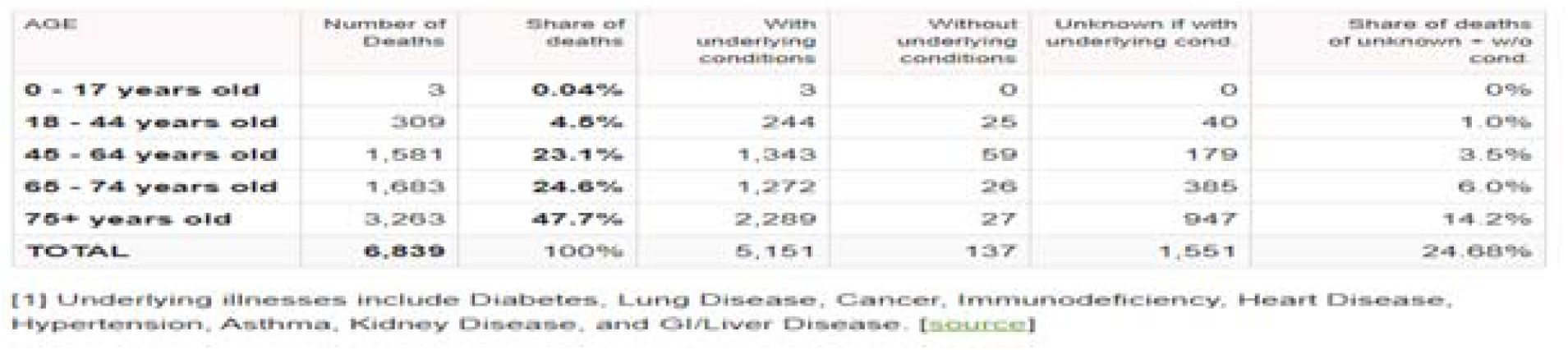
Proportion of people with COVID-19 by age distribution

**Table-2:** Proportion of people affected by COVID-19 by gender distribution

| SEX | Deaths | Share of Deaths | With underlying conditions | Share within this category | Without underlying conditions | Share within this category | Unknown if with cond. | Share within this category |
| --- | --- | --- | --- | --- | --- | --- | --- | --- |
| **Male** | 4,095 | **61.8%** | 3,087 | 62.2% | 96 | 72.2% | 912 | 59.5% |
| **Female** | 2,530 | **38.2%** | 1,873 | 37.8% | 37 | 27.8% | 620 | 40.5% |

As of 8^th^ of May, 2020, the number of cases in Ethiopia was 194 since the first case was reported by 13^th^ of March, 2020.

## 2. Objective

The aim of this review was to characterize and summarize the global COVID-19 pandemic situation and recommend potential mitigation strategies in the context of Ethiopia.

## 3. Methods

### 3.1. Literatures search

The search strategy aims to find both published and unpublished studies. A three-step search strategy was utilized in this review; ***Initial search*** of Pub Med central and Google scholar was undertaken followed by analysis of the text words; COVID19,SARS-CoV-2, Global lessons, Pandemic, A ***second search*** using all identified keywords including Epidemiology, update,COVID19 ***thirdly***, the reference list of all identified reports and articles were searched for additional studies finally Studies published in English insert language was considered for inclusion in this review.

### 3.2. Inclusion and Exclusion criteria

Literatures published as of January 1, 2020 that meet the key words were included in this review. Basic concepts including clinical trials related to vaccine development and treatment were excluded.

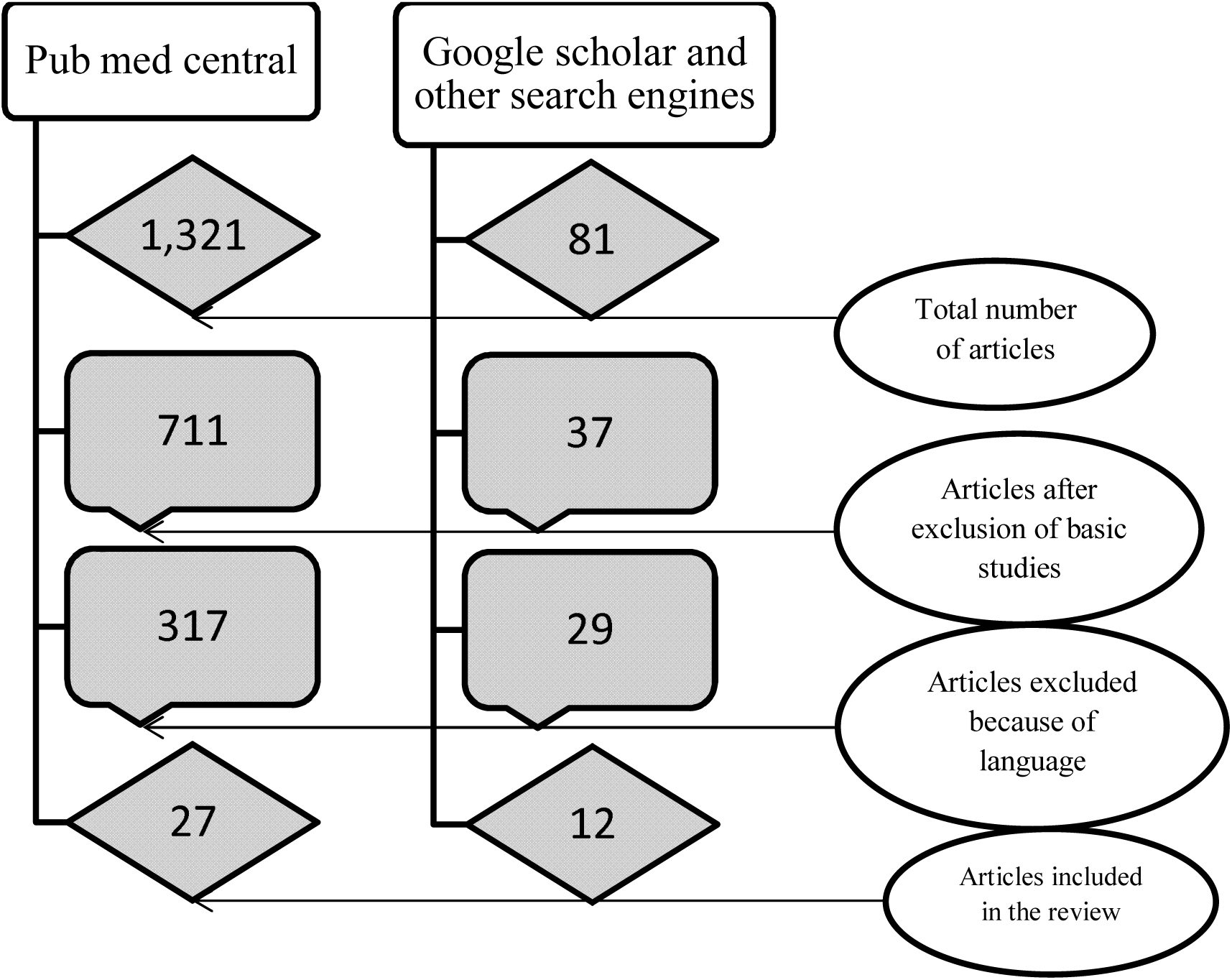

### 3.3. Data extraction and management

The following data was abstracted to a Microsoft Excel spreadsheet: Author, Journal, Year; Setting: Country, number of study participants, design.

### 3.4. Data collection and analysis

Qualitative data was extracted from papers included in the review. The data extracted include specific details about countries experience in responding COVID-19. Finally, textual data were extracted from papers included in the review.

## 4. Result

### 4.1. Global Mitigation strategies

Public health and social measures are measures or actions by individuals, institutions, communities, local and national governments and international bodies to slow or stop the spread of COVID-19. These measures to reduce transmission of COVID-19 include individual, community and environmental measures, detecting and isolating cases, contact tracing and quarantine, social and physical distancing measures including for mass gatherings, international travel measures, and vaccines and treatments^11^. While vaccines and specific medications are not yet available for COVID-19, other public health and social measures play an essential role in reducing the number of infections and saving lives^2^.

Social and physical distancing measures aim to slow the spread of disease by stopping chains of transmission of COVID-19 and preventing new ones from appearing. These measures secure physical distance between people and reduce contact with contaminated surfaces, while encouraging and sustaining virtual social connection within families and communities. Locally adaptable and regularly updated measures for the general public include introducing flexible work arrangements such as teleworking, distance learning, reducing and avoiding crowding, closure of non-essential facilities and services, shielding and protection for vulnerable groups, local or national movement restrictions and staying-at home measures, and coordinated reorganization of health care and social services networks to protect hospitals^8,12^. The measures are used in conjunction with individual protective measures against COVID-19 such as frequent hand washing and cough etiquette^8^.

All public health measures to stop disease spread can be balanced with adaptive strategies to encourage community resilience and social connection, protect incomes and secure the food supply^13^. Countries should balance the possible benefits and negative consequences of each intervention and deploy strategies to encourage community engagement, gain trust and limit social or economic harm. There are many strategies that can support community resilience and mental health, protect access to essential goods and services, and limit the economic impact of stay at home measures where these are deemed necessary. For example, organizing work sites to ensure physical distance between persons, such as staggering shifts over time, or converting onsite service to home delivery may help to keep more businesses open^1^. Tele-working and tele schooling strategies in different contexts demonstrate innovation and the role of technology in supporting business continuity and sustaining social connection within families and communities. In general, while the need for a continues data driven decision at all level is critical, implementation of distancing measures should also aim to sustain personal and professional community connections by virtual means and technology, including widely accessible means such as radio and mobile phones.

Alongside all these measures remains there is the critical to test all suspected cases of COVID-19 wherever possible, promptly isolate cases, trace contacts to the widest extent possible, and ensure quarantine of contacts for the duration of the incubation period. This goes for any context or level of spread of the pandemic in a country, in order to deepen the benefits of social measures. Social measures should make the task of contact tracing much easier as the number of contacts rapidly dwindles and eventually the number of cases declines as well. As social measures are lifted, it is essential to continue to strengthen case-finding, isolation for COVID-19 cases and quarantine of contacts, in order to respond to resurgent or imported cases. Coordinated reorganization of health and social services is essential to assess and test persons rapidly, treat patients effectively, and protect hospitals and health personnel^14^.

WHO has described four levels of COVID-19 transmission which are countries or local areas with: No cases reported, Sporadic cases, Clusters of cases (grouped in place and time), or Community transmission^15^. Countries are putting in place a range of public health and social measures in different combinations and at varying times in the local evolution of the COVID-19 pandemic. The alignment of public health measures to levels of transmission in a community is not fixed in stone. Countries may wish to specify which measures are to be taken at each level and review the situation regularly. A package of measures may be applied at local, regional or national level and adjusted as needed, considering aspects such as culture, living environments, terrain and access to needed resources. Essential services should remain operational and governments should put in place social and economic policies to limit the longer term economic impact, support community resilience and enable rapid recovery. Most importantly, the ultimate aim is to ‘walk back’ community transmission to clusters, sporadic cases, and down to no cases at all, and to begin gradually lifting social measures as soon as it is safe to do so.

To be effective, public health measures must be implemented with the full engagement of all members of society, including communities and professional groups. All measures should be accompanied with clear, accessible and regular risk communication to explain the response strategy and enable people to make informed decisions to protect themselves and help achieve the public health goal of ending the outbreak.

### 4.2. Global lessons in curbing COVID-19 pandemic Universal Safety Precautions

Washing hands is the first line of defense against viruses such as coronavirus^16^. Fear of the public for COVID-19 has significantly contributed towards maintaining the personal hygiene of the individuals. Practicing good hygienic measures in hospitals, schools and other public places could drastically reduce the spread and thereby eliminate new cases^17^.

Governments and other organizations have succeeded in promoting the universal safety precautions such as washing hands, covering nose and mouth while coughing and sneezing, use of sanitizers, use of face masks, avoiding contact of fingers with mouth, nose and eyes, and social distancing techniques to a remarkable extent. Several cities and states across the world are currently short of hand sanitizers and face masks, which show the public’s interest in acquiring the universal precautions to a remarkable extent^18^.

Furthermore, in many countries, efforts are in place to demonstrate proper hand washing and mask usage techniques. Social distancing is a sage practice and an obvious action to be followed during outbreaks for preventing the spread of disease by confining the interaction of individuals and groups^16,19–23^.

Unfortunately, in deferent countries, social distancing measures were not in place until it was too late; as a result, hospitals were filled to capacity, and a rapid transmission was observed, which had led to a steep spike in newer infections. As a result, with the goal of flattening the curve, the Chinese health authorities have implemented non-pharmacological measures such as social distancing, which showed a significant impact in limiting the spread^19^.

A recent modeling study has predicted that if social distancing measures were implemented one week, two weeks and three weeks earlier in China, it could have reduced the number of new cases by 66%, 86%, and 95%, respectively^24^. Furthermore, a New York Times model for the USA has suggested that aggressive social distancing measures could reduce COVID-19 cases from a possible peak of 9 million to 513, 000 and cumulative deaths from 982 000 to 51, 000 over the next few months. Similarly, by adopting the community mitigation measures such as social distancing, countries like South Korea, which experienced a severe outbreak in its initial days, is now remarkably declining its epidemic curve^4,17,18^. Even in USA, if the country provided the social distancing guideline a week and two weeks before the date that was declared it will reduce the death by 60% and 90% respectively^6^.

#### Preparedness among Government Bodies

Consequences of COVID-19 have prepared the government bodies in planning and implementing various measures around the world in better assisting their communities during the disease crisis. Almost every country is doing its best to keep the disease at bay to avoid repeating the nightmares of SARS and MERS.

For instance, China enforcing “round the clock closed management” system, Italy declaring the “red zone” alert, France announcing “nationwide ban on gatherings,” and the USA implementing “containment areas” would be few mentions of what different countries are doing in ceasing the spread^3^.

In addition, to avoid panic and misinformation, measures are in place in enhancing the transparency between government bodies and the public, which can help in educating citizens on the risks of transmission. So far, the experiences of COVID-19 are raw, visceral, and could be considered a rehearsal. This pandemic has clearly made governments know about their strengths and weaknesses regarding their health care systems in responding to outbreaks.

For example, being one of the most leading countries in health care, Italy is unsuccessful in limiting the spread of COVID-19; while, countries like Singapore, Hong Kong, and Taiwan have been hailed for the measures taken to combat the disease and succeeded in keeping their morbidity and mortality rates lower, despite their stronger links to China^4^.

Of note, Taiwan, a country on China’s doorstep, managed to contain COVID-19 by building its public health infrastructure that could be launched during an emergency crisis. They also established a Central Epidemic Command Center that responds to epidemics, biological pathogens, bioterrorism, and medical emergencies^24^.

Lessons Learned by Health Care Sectors This unexpected catastrophe is compelling the health care sectors in tackling the situations. Preparation is a key, and it should be a major lesson to be learned from COVID-19. Hospitals in the USA are implementing disaster readiness and just in case scenario plans, as hospitals will not be notified in advance by a possible COVID-19 case^14,25^.

Also, they are stocking up on equipment and personal protective supplies, including gowns, eye protection and masks. While the risk of COVID-19 to the public is on the rise day by day, hospitals are not taking chances and are doing drills for worst case scenarios. Thus, they are getting ready by setting up quarantine centers, preparing for the extra beds in accommodating the patients, facilitating for infection control, ordering more medical supplies in advance, and organizing cross departmental emergency response committees^26^.

China has built two hospitals that have around 30 intensive care units and hundreds of beds in a span of few days to combat the fast spread of the virus; it would be a prime example that is worth mentioning. However, this is not the case in certain countries, especially the UK, where there is a shortage of general practitioners and hospitals are underequipped. Furthermore, the UK also lacks enough beds and personal protective equipment for health care staff while dealing with the patients during this crisis^13,27^.

### 4.3. Potential strategies in Ethiopian context in combating COVID-19

#### Individual, community and environmental measures

One of the most feasible and important individual based preventive measure is proper hand washing. For which, wide use of mainstream and social media for improving knowledge, attitude and practice on hand hygiene, use of masks and sanitizers. It is also important to ensure availability of supplies which include masks, sanitizer, sops, and other supplies. Use of health extension workers with assignment of specific catchment population to regularly monitor the health of the community at household level may be important. Mobilizing the youth for the prevention and control is also vital part.

#### Detecting and isolating cases

Many countries are facing critical challenge in this regard mainly because of the limited of access to testing. Hence, it is crucial to consider innovative strategies since it will not be feasible to address the community using the available PCR based testing method in resource limited setting like Ethiopia. It is therefore to consider validation and availability of rapid diagnostic test kits for COVID-19 testing. The entries for identifying cases are health facilities. Hence, it is crucial to update the case definitions of COVID-19 for early identification of cased and pick cases with mild symptom. For instance, the US CDC revised the previous case definition and currently; Cough, Shortness of breath, Fever, Chills, Muscle pain, Sore throat or New loss of taste or smell are considered as symptoms for COVID-19 and individuals visiting health facilities with either of these symptoms will be tested for COVI-19. In Ethiopian context, health extension workers could be assigned to the community with a specific catchment population to monitor the health of the community for COVID-19 in a regular basis using the case definition in facilitating identifying the cases. It could also be crucial to use the long standing effective strategy on HIV to adapt to COVID-19 mitigation.

#### Contact tracing and quarantine

Countries experience in this regard revealed that, there is a 24/7 workforce for contact tracing and there is a dedicated space for quarantine. However, quarantine for 14 days is becoming the critical issue since some reports from WHO and other scientific reports revealed that the incubation period of COVID-19 could go to 24 days. Hence, it could be important to revise the date of quarantine.

#### Social and physical distancing measures including for mass gatherings

Social and physical distancing is the most effective but the most challenging measure so far. Proper implementation of social and physical distancing can curb the pandemic to 95%. The global challenge here is the time when to order and its level of implementation. From experience, countries that strictly implement social and physical distancing contain the pandemic at the earliest and the earlier the order and implementation, the minimal disaster due to the pandemic. The critical challenge here is coordination, community engagement and public awareness. Use of different social media to aware the public is important. In the context of the country, it could be important to consider active involvement of religious leaders, mobilizing youth and use of influential people including artists. From countries experience, law enforcement is the important component.

Multisectoral approach (MSA) refers to deliberate collaboration among various stakeholder groups (e.g., government, civil society, and private sector) and sectors (e.g., health, environment, and economy) to jointly mitigate COVID-19 is crucial. By engaging multiple sectors, partners can leverage knowledge, expertise, reach, and resources, benefiting from their combined and varied strengths as they work toward the shared goal of producing better health outcomes. Improving public health (PH) is challenging because of the size of its population and wide variation in geography. MSA help in addressing identified health issues in focused way as it helps in pooling the resources and formulating the common objectives. One of the major advantages is optimization of usage of resources by avoiding duplication of inputs and activities which tremendously improve program effectiveness and efficiency. Willingness at the leadership and mandate at the policy level are necessary to plan and execute the successful multisectoral coordination. All the major stakeholders require sharing the common vision and perspective. Developing institutional mechanism is utmost requirement as it will standardize the processes of intersectoral coordination (ISC). Creation of PH cadre is strategic move to meet the major health challenges being faced by the health system, and it would be anchor of establishing systematic ISC. There are many national and international examples of MSA applications such as for malaria elimination, tobacco control and HIV/AIDS prevention. Promotion of MSA within the health system and with other ministries is seen as an important measure for effective implementation and improving efficiency.

#### International travel measures

Many countries in this regard restrict international travel from the highly affected countries and countries have strong monitoring strategy for international travelers. The good example is in Ethiopia, there is 14 days mandatory quarantine for international travelers. However, 14 days quarantine might not be enough since different reports revealed up to 24 days incubation period for COVID-19.

#### Vaccines and treatments

The global scientific community is working round clock to gate safe vaccine and treatment. African countries harbor many indigenous plants which could be candidate for potential treatment and it would be a great opportunity, while strictly following the international standards, to encourage testing those for potential treatment.

## 5. Conclusion

In a nutshell, this pandemic has reiterated the importance of a saying “prevention is better than cure” and has psychologically prepared mankind to battle and combat this pandemic. It has also revealed weak points in how we think about health and prepare for the disease. Coronavirus is not only a curse, but also a chance to improve our facilities and health care infrastructure and, above all, to learn how to be more ready for the next emergency crises.

This new virus outbreak has challenged the economic, medical and public health infrastructure of countries. More so, future outbreaks of viruses and pathogens of zoonotic origin are likely to continue. Therefore, apart from curbing this outbreak, efforts should be made to devise comprehensive measures to prevent future outbreaks of zoonotic origin.

## Data Availability

Since this is systematic review, the is no raw data

## Notes

### Competing Interest Statement

The authors have declared no competing interest.

### Funding Statement

No finding for this review

### Author Declarations

Since this is systematic review, Ethical review was not applicable

## References

1. Jennifer, M. Characteristics of and Important Lessons From the Coronavirus Disease 2019 (COVID-19) Outbreak in China Summary of a Report of 72 314 Cases From the Chinese Center for Disease Control, and Prevention. 323, (2020).

2. Wang, J. & Wang, Z. Strengths, Weaknesses, Opportunities and Threats (SWOT) Analysis of China ‘ s Prevention and Control Strategy for the COVID-19 Epidemic. 2019, (2020).

3. Sohrabi, C. et al. World Health Organization declares global emergency□: A review of the 2019 novel coronavirus (COVID-19). Int. J. Surg. 76, 71–76 (2020).

4. Report, S. Coronavirus disease 2019 (COVID-19). 2019, (2020).

5. Singhal, T. A Review of Coronavirus Disease-2019 (COVID-19). 87, 281–286 (2020).

6. Salzberger, B., Glück, T. & Ehrenstein, B. Successful containment of COVID - 19□: the WHO □ Report on the COVID - 19 outbreak in China. Infection 19–21 (2020). doi:10.1007/s15010-020-01409-4

7. Harapan, H., Itoh, N., Yufika, A., Winardi, W. & Keam, S. Coronavirus disease 2019 (COVID-19): A literature review. (2020).

8. Freedman, D. O. Isolation, quarantine, social distancing and community containment□: pivotal role for old-style public health measures in the novel coronavirus (2019-nCoV) outbreak. 1–4 (2020). doi:10.1093/jtm/taaa020

9. Wuhan novel coronavirus (COVID-19): why global control is challenging? 19–21 (2020).

10. Ferioli, M. et al. FRONTIERS IN CLINICAL PRACTICE Protecting healthcare workers from SARS-CoV-2 infection 2020: practical indications. (2020). doi:10.1183/16000617.0068-2020

11. Nicola, M. et al. Evidence based management guideline for the COVID-19 pandemic - Review article. Int. J. Surg. 77, 206–216 (2020).

12. Article, R. COVID-19: Prevention and control measures in community. 571–577 (2020). doi:10.3906/sag-2004-146

13. Mayr, V. et al. measures to control COVID-19: a rapid review (Review). (2020). doi:10.1002/14651858.CD013574. www.cochranelibrary.com

14. Wang, L., Wang, Y., Ye, D. & Liu, Q. Review of the 2019 novel coronavirus (SARS-CoV-2) based on current evidence. Int. J. Antimicrob. Agents 105948 (2020). doi:10.1016/j.ijantimicag.2020.105948

15. Search, G. Coronavirus COVID-19 situation reports - World Health Organization. 5–7 (2020).

16. Adhikari, S. P. et al. Epidemiology, causes, clinical manifestation and diagnosis, prevention and control of coronavirus disease (COVID-19) during the early outbreak period: a scoping review. 1–12 (2020).

17. Watkins, J. Preventing a covid-19 pandemic We need to think beyond containment. 810, 1–2 (2020).

18. Dong-hoon, K. & Canizales-quinteros, S. Obesidad, tejido adiposo y cirugía bariátrica. 77, 3–14 (2020).

19. Th, I. N. D. E. P. Can China’s COVID-19 strategy work elsewhere?

20. Surveillances, V. The Epidemiological Characteristics of an Outbreak of 2019 Novel Coronavirus Diseases (COVID-19) — China, 2020. 2, 113–122 (2020).

21. Sjödin, H., Wilder-smith, A., Osman, S., Farooq, Z. & Rocklöv, J. Only strict quarantine measures can curb the coronavirus disease (COVID-19) outbreak in Italy, 2020. 1–6 (2020).

22. Ebenso, B. & Otu, A. Correspondence Can Nigeria contain the COVID-19 outbreak using lessons from. Lancet Glob. Heal. 30101 (2020). doi:10.1016/S2214-109X(20)30101-7

23. How will country-based mitigation measures influence the course of the COVID-19 epidemic? (2020).

24. Gudi, S. K. & Tiwari, K. K. Preparedness and Lessons Learned from the Novel Coronavirus Disease. 11, 108–112 (2020).

25. Tian, S. et al. Characteristics of COVID-19 infection in Beijing. J. Infect. 80, 401–406 (2020).

26. Yang, J. et al. Prevalence of comorbidities and its effects in patients infected with SARS-CoV-2D: a systematic review and meta-analysis. 94, 91–95 (2020).

27. Handling the COVID-19 pandemic in the oncological setting _ Elsevier Enhanced Reader.pdf.

